# Evaluation of inverse treatment planning for Gamma Knife radiosurgery using fMRI brain activation maps as organs at risk

**DOI:** 10.1101/2022.12.12.22283334

**Authors:** David Abramian, Ida Blystad, Anders Eklund

## Abstract

Stereotactic radiosurgery (SRS) can be an effective primary or adjuvant treatment option for intracranial tumors. However, it carries risks of various radiation toxicities, including radionecrosis and functional deficits. Current SRS inverse planning algorithms allow efficient inclusion of organs at risk (OARs) in the treatment planning process, which will be spared by setting a maximum radiation dose. In this work we propose using activation maps from functional MRI to map the eloquent regions of the brain and define functional OARs. We evaluate the effects of these functional OARs for Elekta Leksell Gamma Knife SRS inverse treatment planning on open data MRI from 5 subjects. Our results show that fMRI-derived functional OARs can effectively be used to reduce the radiation dose incident on the eloquent brain regions, while maintaining acceptable treatment planning metrics on the tumor targets.

## 1. Introduction

Brain tumors compose about 2% of new cancer diagnoses, affecting some 300 000 subjects globally each year (Leece et al., 2017). Although not the most prevalent cancer type, brain tumors are prone to complicated and challenging treatment procedures, which often combine surgical resection, radiotherapy, and chemotherapy, with a high morbidity and low survival rate for the patients. Stereotactic radiosurgery (SRS) can be an efficient treatment option for small primary brain tumors and metastases. However, treatment planning requires considerable time, as it involves collecting MR and/or CT images, segmenting the tumor and organs at risk (OARs), and the generation of treatment plans. Furthermore, OARs typically include various anatomical landmarks, but eloquent regions of the brain are rarely used for this purpose. In this work, we describe a workflow for efficiently incorporating functional OARs (fOARs), in the form of brain activation maps derived from functional MRI (fMRI), into Gamma Knife SRS treatment planning in order to minimize the radiation dose incident on eloquent brain areas.

### 1.1. Stereotactic radiosurgery

Stereotactic radiosurgery is an image-guided non-invasive surgical approach which relies on focused beams of radiation to ablate tissue. Originally developed to treat various benign intracranial conditions, it has since seen its use extended to malignant conditions, both intracranial and elsewhere in the body.

Since its inception, SRS has been adopted within the fields of neurosurgery and radiation oncology. It has become one of the treatments of choice for multiple intracranial conditions and malignancies, such as arteriovenous malformations, vestibular schwannomas, cavernous sinus meningiomas, recurrent or residual pituitary tumors, metastatic tumors, and trigeminal neuralgia (Lunsford & Sheehan, 2009). It is also increasingly being applied for the treatment of high- and low-grade gliomas (Fetcko et al., 2017; Bokstein et al., 2016; Skeie et al., 2012; Weintraub et al., 2012) and meningiomas (Ruge et al., 2021; Pinzi et al., 2017; Han et al., 2017).

### 1.2. Handling risk organs

Despite the precise dose delivery and sharp dose fall-off achievable with radiosurgery, patients are still at risk of suffering various radiation-induced toxicities. Complications associated with SRS for vestibular schwannoma include facial neuropathy, hearing loss and damage to the trigeminal nerve (Kondziolka et al., 1998). High grade glioma treatment complications include radionecrosis, cranial nerve palsy, paralysis, seizures, CNS hemorrhage, and stroke (Fetcko et al., 2017). A large scale meta-analysis of published research on Gamma Knife radiosurgery for arteriovenous malformations found that 34 % of the patients developed radiation-induced changes, with 8 % developing neurological symptoms and 3 % experiencing permanent neurologic deterioration (China et al., 2022).

In order to mitigate potential radiation-induced functional deficits to patients treated with SRS, it is common to segment various organs at risk (OARs) during the treatment planning process, and keep the dose incident on them within established limits by shaping the radiation dose delivered to the targets. For SRS of the brain, these OARs typically include anatomical regions such as the brain stem, the hippocampus, the optic nerves, and the cochlea.

### 1.3. Using risk organs from functional MRI

Since its development in 1992, fMRI has been used to study brain activity and brain connectivity (Logothetis, 2008). It has become very popular for both research and clinical applications, as it is based on a natural contrast mechanism that is part of the human physiology.

fOARs derived from fMRI have been previously used to spare the eloquent brain regions in radiotherapy (Narayana et al., 2007; Kovács et al., 2011; Wang et al., 2015), as well as in SRS with LINAC (Liu et al., 2000) and CyberKnife (Stancanello et al., 2007; Pantelis et al., 2010; Conti et al., 2013; De Martin et al., 2017), but, to the best of our knowledge, not with Gamma Knife. One reason for this could be the availability of inverse planning solutions. While treatment plans with multiple anatomical and/or functional OARs can be designed with manual forward planning, the process can be too time-consuming, depending on the number and size of the targets and the OARs, as well as the distances between them. Modern inverse planning methods for radiation delivery (Sjölund et al., 2019; Paddick et al., 2020; Zeverino et al., 2019) greatly simplify the incorporation of fOARs into the treatment planning routine, using automatic optimization to generate treatment plans that consider multiple OARs within minutes. The recent introduction of the Elekta Gamma Knife Lightning treatment planner (Sjölund et al., 2019) (henceforth referred to as Lightning) enables the creation of complex treatment plans with multiple OARs for Gamma Knife SRS.

In this work we have developed a pipeline for analyzing fMRI data to create fOARs, and converting the anatomical images and fOARs into DICOM objects that can be imported into the Elekta Leksell GammaPlan software (henceforth referred to as GammaPlan, see Figure 1), to be used with the Lightning inverse planner add-on. We used this pipeline to generate retrospective treatment plans for 5 brain tumor patients and evaluated the effects of including fOARs in the treatment planning process with the Lightning planner.

**Figure 1:**
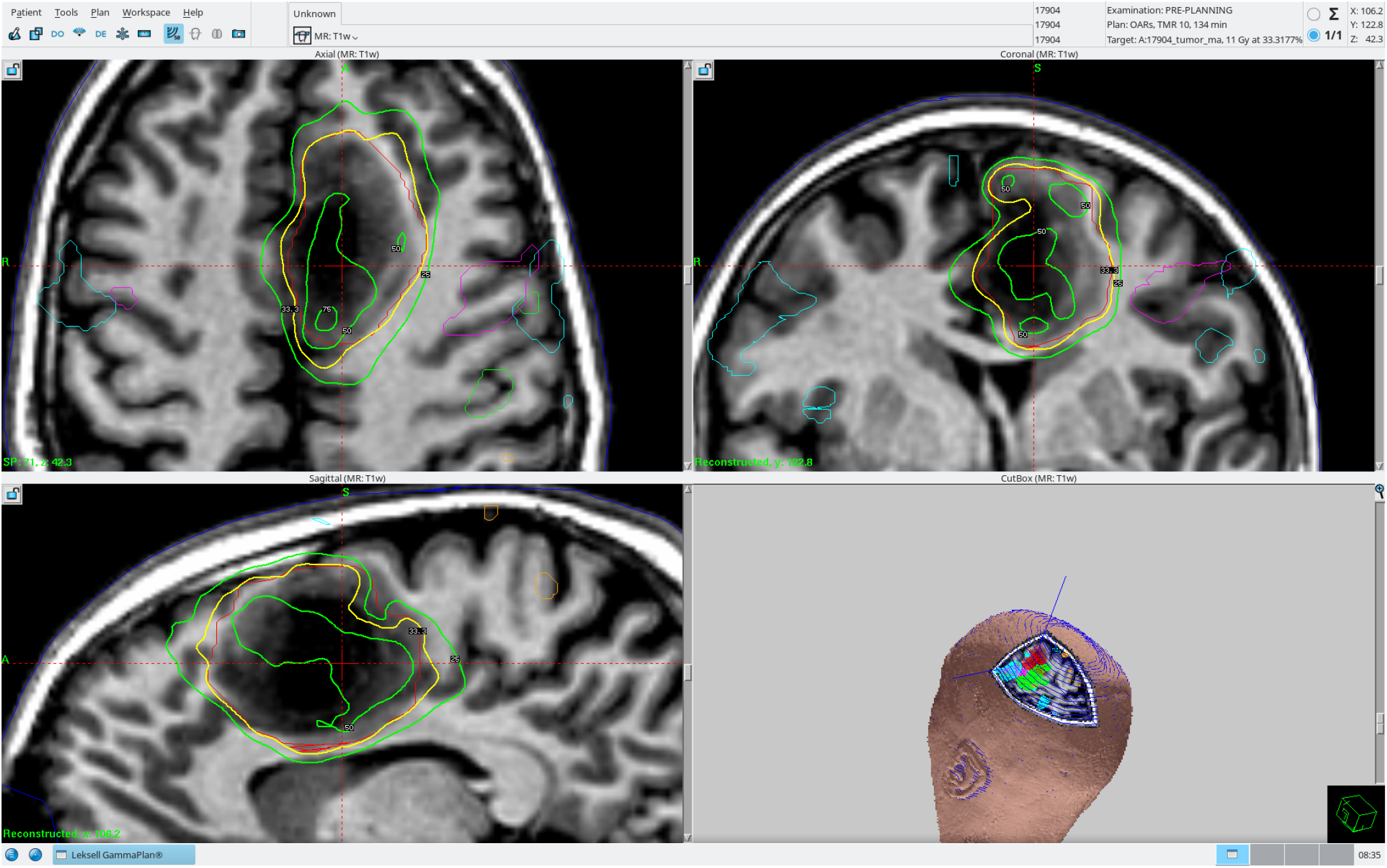
An example of treatment planning in GammaPlan after loading the anatomical images, the tumor mask and the risk organs obtained from fMRI. Red contours represent the tumor mask. Thick yellow and green contours represent the 11 Gy and 4 Gy isolevels, respectively, corresponding to the target tumor irradiation dose and the maximum dose constraint on fOARs, respectively. Thin green, orange, cyan, and magenta contours represent fOARs defined from the hand, foot, lips and verb repetition fMRI conditions, respectively.

## 2. Data

We used an open MRI dataset of brain tumor patients to carry out our experiments (Pernet et al., 2016). The dataset contains structural (T1w, T2w, DWI) and functional MRI data from 22 patients with various types of brain tumors located close to the eloquent regions of the brain. Functional imaging protocols include a motor task consisting of three conditions (finger tapping, foot flexing, lip pursing), a covert verb generation task (mapping of Broca area and supplementary motor area), an overt word repetition task (mapping of Wernicke area), as well as resting state fMRI. The employed fMRI protocol has previously been shown to provide reliable singe-subject activations on control subjects (Gorgolewski et al., 2013). Due to medical considerations, not all fMRI datasets are available for all patients. While we are aware of larger open brain tumor datasets, such as the BraTS dataset (Menze et al., 2014), including 1251 subjects in the training set of the 2021 version, they do not generally include fMRI data, and are thus unsuitable for this work. Due to limitations in the suitable radiation doses and treatment times for a patient, SRS is mainly applied on relatively small pathologies. Because of this, we only used data from patients with tumors smaller than 40 cubic centimeters, which excluded 17 of the 22 patients in the dataset. Table 1 presents information on the remaining 5 subjects that were used in our analyses.

**Table 1:** An overview of the patients analyzed in this study. Patients with tumors larger than 40 cubic centimeters were excluded from the original Pernet et al. (2016) dataset, since SRS is normally not used for such patients. Not all fMRI datasets are available for all patients. M = motor, V = verb generation, W = word repetition, R = resting state.

| Subject ID | Pathology | Tumor location | Tumor size | Available fMRI data |
| --- | --- | --- | --- | --- |
| 17904 | Oligodendroglioma type II | Left supplementary motor area | 35.92 cm<sup>3</sup> | M V R |
| 18582 | Astrocytoma type II | Wernicke area | 10.34 cm<sup>3</sup> | M V W |
| 18975 | Glioblastoma multiforme | Left primary motor area | 13.24 cm<sup>3</sup> | M V W R |
| 19015 | Glioblastoma multiforme | Left supplementary motor area | 37.40 cm<sup>3</sup> | M V W |
| 19849 | Not available | Right primary motor area | 9.66 cm<sup>3</sup> | M R |

For each subject we made use of the T1w volume as reference for treatment planning, and the task fMRI data was used to obtain functional OARs (fOARs). Table 2 presents some relevant acquisition parameters for these modalities. While the dataset includes tumor masks, these were generated using a semi-automatic procedure. To ensure the accurate localization of the tumors, co-author IB, an experienced neuroradiologist, manually annotated the tumor for each of the patients used in our analyses.

**Table 2:** Acquisition parameters for the Pernet et al. (2016) dataset.

|  | T1w | Motor | Verb generation | Word repetition |
| --- | --- | --- | --- | --- |
| Voxel size [mm] | 1 × 1.3 × 1 | 4 × 4 × 4 | 4 × 4 × 4 | 4 × 4 × 4 |
| Dimensions [vox] | 256 × 156 × 256 | 64 × 64 × 30 | 64 × 64 × 30 | 64 × 64 × 30 |
| TR [s] | 10 | 2.5 | 2.5 | 5 (effective 2.5) |
| Number of volumes | 1 | 184 | 173 | 76 |

## 3. Methods

This section describes the methods used to generate the fMRI activation maps and use them as fOARs for treatment planning in GammaPlan. All data processing and conversion was done using a combination of Bash, Matlab, and Python scripts, which are available online^1^. A diagram of our processing and analysis pipeline is shown in Figure 2.

**Figure 2:**
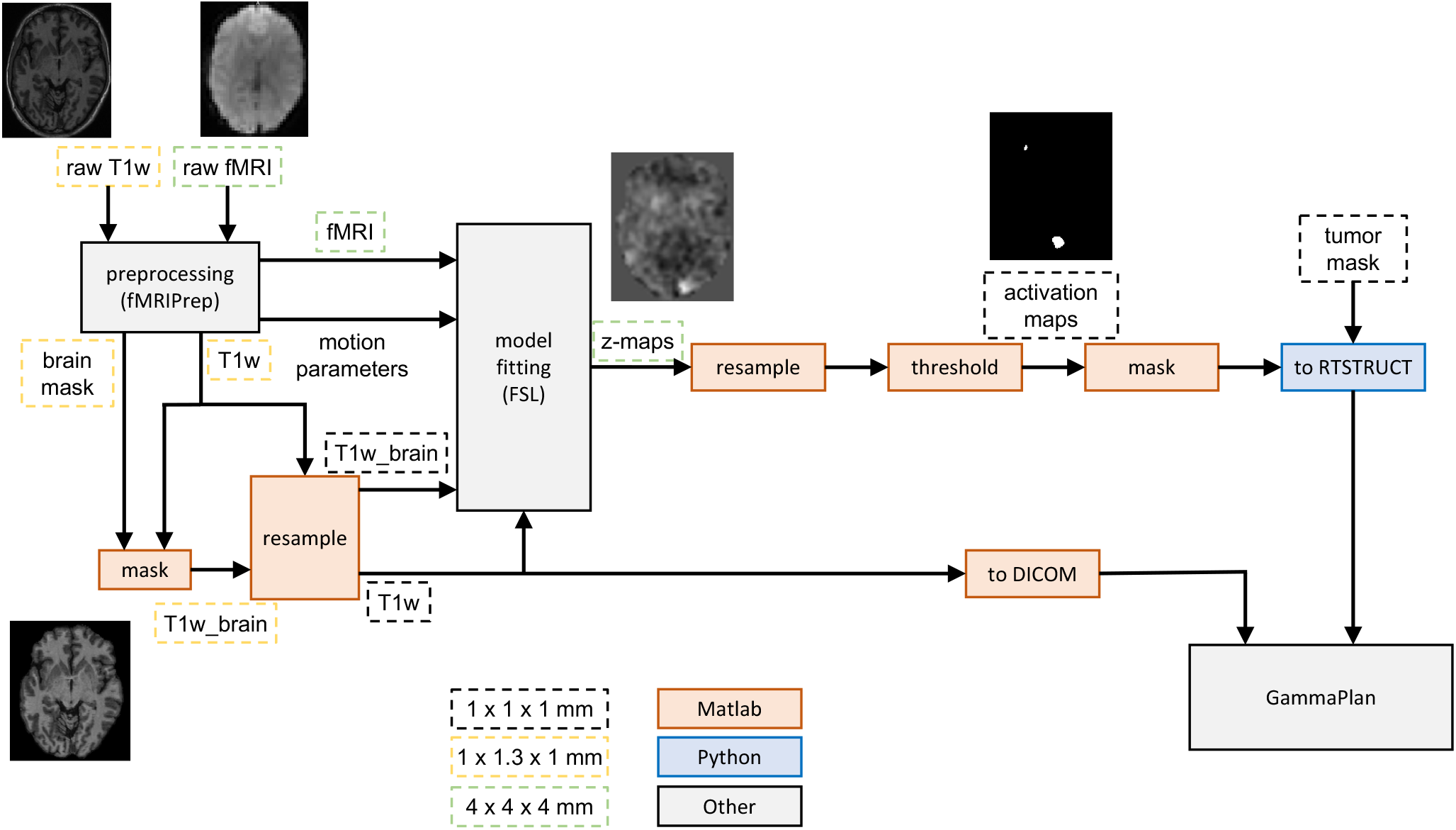
Diagram of the preprocessing and statistical analysis pipeline for fMRI data. The preprocessing (e.g., registration between T1w and fMRI volumes, head motion correction) is done with the fMRIPrep software, with some additional resampling of the volumes. The statistical analysis of the fMRI data is done with the FSL software. A custom Python script is finally used to convert the brain activity maps to DICOM RTSTRUCT, for loading them into GammaPlan.

### 3.1. Preprocessing of MRI data

For each subject, the T1w and fMRI data are available in NIfTI format. The first four volumes of all fMRI sequences are dummy volumes, and were deleted.

We chose to preprocess the anatomical and functional MRI data with the fMRIPrep software (Esteban et al., 2019), which purposefully combines processing steps from multiple MRI software tools to produce a robust and versatile preprocessing pipeline in accordance with best practices. As fMRIPrep requires the data to be in the BIDS format (Gorgolewski et al., 2016), the dataset was first manually converted to BIDS. The preprocessing steps included bias correction of the T1w volume, brain mask extraction, head motion correction of the fMRI volumes, and co-registration of fMRI data to the corresponding T1w anatomical references, among others. A full description of the preprocessing performed by fMRIPrep is given in Appendix A.

We experimented with several different settings for fMRIPrep with and without the inclusion of brain surface extraction with FreeSurfer (Fischl, 2012) for the refinement of the brain mask. Given the substantial increase in processing time from the use of surface extraction (6 hours per subject vs. 1.5 hour per subject) and the minimal differences observed in the final results, we decided not to use surface processing for our experiments. Other fMRIPrep settings used included producing output in the native space of each subject, and using experimental fieldmap-free susceptibility distortion correction for the fMRI data, due to the absence of fieldmaps in the dataset.

Following preprocessing with fMRIPrep, we used the generated brain mask to produce a skull-stripped version of the T1w volume, to be used in FSL. Both versions of the T1w volume were then resampled to 1 mm isotropic resolution and 256 *×* 256 *×* 256 voxels in order to fulfill a requirement from the GammaPlan software, namely that the 2D slices of the reference volume have square pixels. These images were used by co-author IB to produce new tumor segmentations, so these were made in the target resolution.

### 3.2. Activation mapping

We used the FSL 6.0.4 software (Jenkinson et al., 2012) to perform first-level analysis of the fMRI data. Preprocessing steps were disabled, as they were already carried out by fMRIPrep, with the exception of spatial smoothing. Due to the importance that smoothing has for the detection power and spatial specificity of found activations, we decided to test Gaussian filters of two different sizes for our analyses, with full width at half maximum (FWHM) sizes of 4 mm and 6 mm, respectively. The GLM design matrices included an activity regressor for each task, the temporal derivative of each activity regressor, and a standard set of six head motion regressors estimated by fMRIPrep.

Prior to thresholding the Z-static maps, these were resampled to the same resolution of the T1w reference volume, as necessary setup for the later conversion of the activation maps into DICOM RTSTRUCT formatted data (see Section 3.4). The resampled Z-statistic maps were then thresholded at an arbitrary voxel level of *Z* = 5, with no cluster extent threshold, to produce binary activation maps. While arbitrary, this threshold provides relatively strong familywise error control, corresponding to Bonferroni correction with *α* between 0.006 and 0.008 for the various subjects and tasks. This activation mapping scheme did not produce any activations for the word repetition task for any subject, so this task was disregarded in the remainder of our analyses.

The arbitrary nature of our chosen thresholding scheme was considered appropriate for several reasons. Firstly, there is currently no consensus on how to best threshold brain activation maps for single subject fMRI analysis. This is especially true for clinical applications, where the relative importance of false positives and false negatives is different to that in research (Silva et al., 2018). Furthermore, ad hoc thresholds are routinely used in clinical fMRI, where the operator manually adjusts the thresholds until a satisfactory activation map had been achieved (Stippich et al., 2022), or sets them in accordance with the relative ease of detecting activations for different tasks (Stancanello et al., 2007). As our work is not focused on settling these questions but on demonstrating the feasibility of generating and using fOARs for Gamma Knife treatment planning, we chose an arbitrary thresholding level with strong control of familywise error rates.

### 3.3. Masking of activation maps

For several subjects and tasks, fMRI activations were present inside or in close proximity to the tumor mask. It is unclear whether these activations represent true brain activity or they are artifactual, potentially caused by tumor-induced neurovascular uncoupling (Bogomolny et al., 2004; Silva et al., 2018). However, as the GammaPlan inverse planner treats the maximum dose on OARs as a hard optimization constraint, it prioritizes reducing the radiation dose incident on OARs over fulfilling the irradiation requirements on the tumor mask, resulting in poor quality treatment plans. In order to address this issue, we created three additional masked versions of each activation map: one where activations found inside the tumor mask were removed, and two where activations were also removed within 2 mm and 4 mm of any tumor voxel, respectively. Thus, for each subject and condition we generated 8 activation maps in total (2 smoothing settings *×* 4 masking settings).

### 3.4. Data conversion to DICOM

As a clinical software, the GammaPlan treatment planning software accepts input data in the DICOM format, ensuring interoperability with other medical devices. However, most fMRI processing and analysis tools require data in the NIfTI format, standard in neuroimaging research, so this is the data format used throughout our pipeline. Therefore, a data conversion step was needed before the brain activation maps could be loaded onto GammaPlan.

The various ROIs, such as the tumor mask and activation maps, need to be provided as DICOM radiotherapy structure sets (RTSTRUCTs) (Law & Liu, 2009), used for the storage and transfer of patient structures. In this format ROIs are represented as collections of contours encompassing the relevant region, with each contour being associated to a reference DICOM object, which in our case are individual T1w axial slices. A single RTSTRUCT file can contain any number of separate ROIs, each of which can be used as a target or risk organ in GammaPlan. Contrastingly, the most common way of representing ROIs in the NIfTI format is as binary volumes, with voxels taking a value of 1 if belonging to the ROI and 0 otherwise.

The T1w reference volume of each subject was split into 256 axial slices, and each of them converted into a DICOM file. The conversion of ROIs from solid 3D regions into collections of 2D contours was done using a custom Python script. The active voxels were first extracted for each axial slice separately, following which the contrours enclosing each connected regions were found automatically using the `find_contour` routine from the `scikit-image` package (Van der Walt et al., 2014), which relies on the marching squares algorithm for finding contours. The sets of contours were converted into RTSTRUCT files using the `pydicom` package (Mason, 2011). Specifically, as RTSTRUCT files can be quite complex, we used the `codify` function to deconstruct an existing RTSTRUCT file and generate the Python code that would recreate it, and altered this code to suit our needs.

### 3.5. Treatment planning

A non-clinical version of GammaPlan^2^ was set up on an HP Z6 G4 workstation with a 10 core Intel Xeon 2.2 GHz CPU, an Nvidia Quadro P2000 GPU, and 96 GB of RAM, in accordance with the required specification for the software, with the exception of the increased RAM (96 GB instead of 32 GB). All analyses were performed with a treatment dose rate of 3.5 Gy/min, corresponding to a new Gamma Knife unit.

For each subject, the set of DICOM files containing the T1w reference volume and the RTSTRUCT files containing the tumor mask and activation map ROIs were transferred to the GammaPlan workstation using a USB drive and imported into the software. Each subject and set of parameters were analyzed in separate examinations within the software. For each such examination, the T1w reference volume and relevant tumor and fOAR ROIs were loaded. In all cases, all of the available activation maps for each subject were used simultaneously, yielding between 80 and 180 risk organs (i.e. individual brain activity clusters, see section 5.3). The skull was segmented automatically in GammaPlan from the T1w volume.

We defined two treatment plans in each examination: one where only the target dose for the tumor is specified, and another were dose limits were also set for each fOAR. The doses incident on the tumor and fOARs were logged in both cases. We used a target dose of 11 Gy and a dose limit of 35 % of the target dose (4 Gy) for all fOARs in all cases.

Optimized treatment plans were generated using the Lightning inverse planning system (Sjölund et al., 2019). It provides two tuning parameters for setting the relative importance of minimizing the dose incident on tissue outside the target and minimizing the beam-on-time, respectively. These were set to their default value of 0.5. The optimization of individual treatment plans took between 20 seconds and 30 minutes.

### 3.6 Exporting GammaPlan results

As treatment plan optimization times are not reported in GammaPlan, we measured them by tracking the timing of changes in on-screen dialog boxes, using motion tracking software running on another computer with a webcam. Furthermore, while it is possible to export complete treatment plans in DICOM format, detailed dose reports for ROIs are only presented in a table to be visualized by the GammaPlan operator, and can not be exported in a machine-readable format on a single-treatment-plan basis. GammaPlan offers the possibility of exporting the full debug logs of the treatment planning computer to external media, which include a complete history of treatment planning logs with detailed statistics in JSON format, but these do not include minimum and mean dose statistics for ROIs other than the targets. In addition, this process is inconvenient for exporting the metrics of individual treatment plans of interest, as it is time-consuming and produces a large number of log files unrelated to this task. Therefore, in order to extract the full treatment planning statistics reported by GammaPlan, we took screenshots of the results tables and exported them as PDF files to a USB drive. We then used custom OCR software written in Matlab to parse the numbers from the tables. The dose statistics for all ROIs that constitute a single fMRI condition were combined.

## 4. Results

For every subject, spatial filter size, and masking setting, we compared the quality of treatment plans with and without the inclusion of fOARs with different masking settings in the optimization process. In total we generated 50 treatment plans (5 subjects *×* 2 smoothing levels *×* (no fOARs + fOARs with 4 masking settings)). Figure 3 shows example treatment plans for every subject and setting tested.

**Figure 3:**
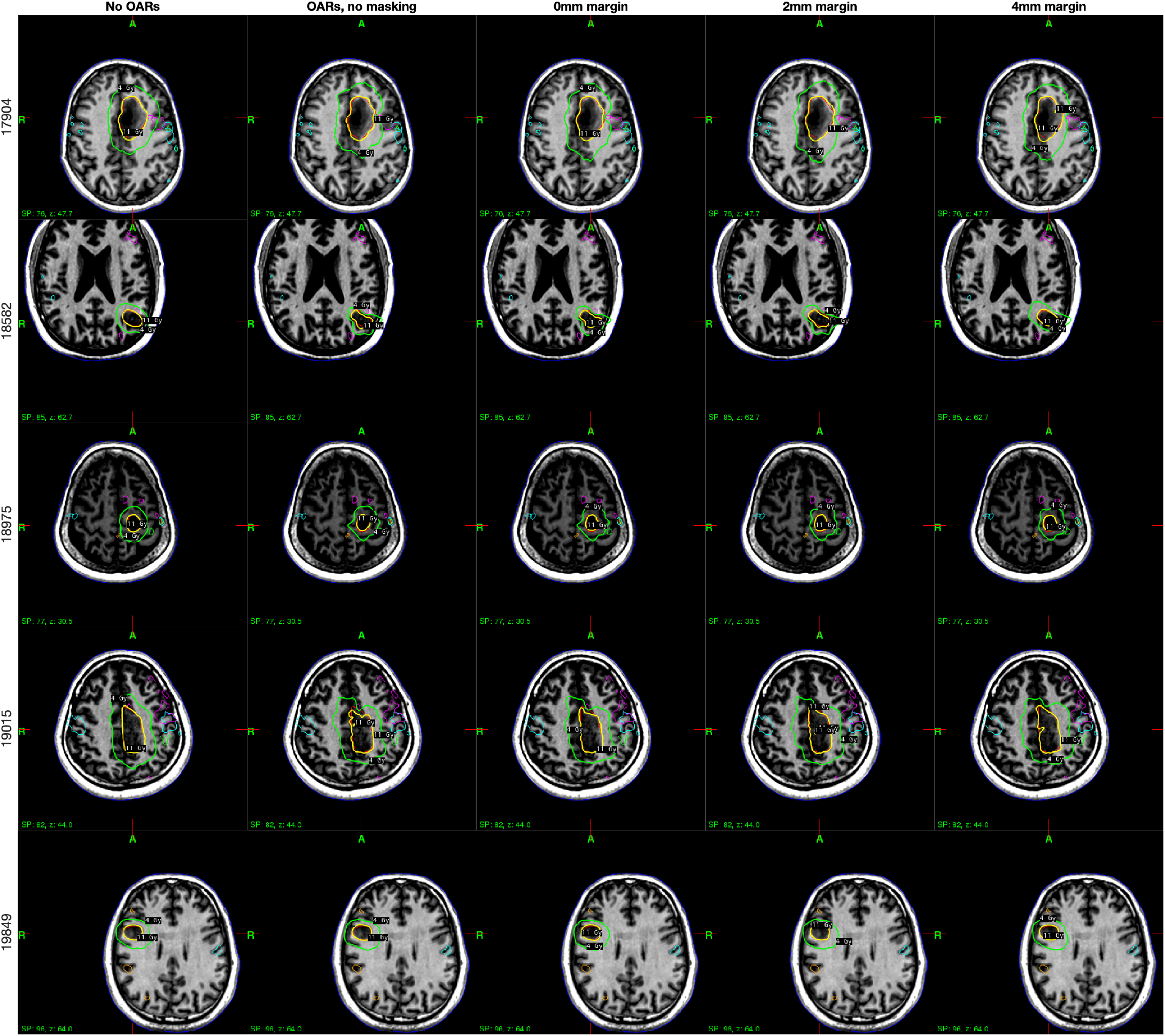
Example treatment plans obtained for all subjects and masking settings, using spatial smoothing with a 4 mm FWHM filter. The first column (“No fOARs”) shows results where no dose limits are set for the fOARs, while the remaining four columns represent the various masking settings employed. Thick yellow and green contours represent the 11 Gy and 4 Gy isolevels, respectively, corresponding to the target tumor irradiation dose and the maximum dose constraint on OARs, respectively. Thin magenta, cyan, orange and green contours represent fOARs defined from each fMRI condition (three from the motor task and one from the verb generation task).

We evaluated the treatment plans on the basis of the metrics reported by GammaPlan. These include minimum, maximum and mean doses for each ROI (tumor mask and fOARs), as well as the following overall treatment plan quality metrics: target coverage, Paddick conformity index (PCI) (Paddick, 2000), gradient index (Paddick & Lippitz, 2006), and treatment time. In addition, we report the optimization time needed to produce each treatment plan. Note that GammaPlan did not report gradient indices for the subjects with larger tumors, so results are only provided for subjects 18582, 18975 and 19849.

### 4.1. Effects of including fOARs

Treatment planning without the inclusion of fOARs results in highly conformal treatment plans, with a target coverage of 1 in all cases, a median PCI of 0.89 (range 0.86 – 0.90), and a median gradient index of 2.66 (range 2.62 – 2.75) where reported. The median minimum, maximum and mean doses delivered to the tumors are 9.9, 20.7 and 15.2 Gy, respectively, reflecting good compliance with the target doses. However, the doses received by the fOARs are in excess of the permissible maximum, with a median maximum dose on fOARs of 6.7 Gy, and reaching doses larger than 10 Gy in several cases.

Incorporating maximum dose constraints for fOARs successfully limits the dose received by them, lowering the median maximum dose on fOARs to 3.9 Gy, i.e., just under the maximum constraint of 4 Gy. In spite of that, the presence of fMRI activations within or in close proximity to the tumor masks results in the inclusion of dose constraints for fOARs having a severe detrimental effect on treatment plan metrics. In this case the median target coverage and PCI are reduced to 0.80 (range 0.60 – 0.97) and 0.70 (range 0.48 – 0.81), respectively, while the median gradient index increased to 2.88 (range 2.74 – 3.19). Furthermore, while the mean tumor doses remained approximately constant, the treatment plans suffer from both hot and cold spots, as indicated by high maximum and low minimum doses, with medians of 2.25 Gy and 32.45 Gy, respectively.

As illustrated by visually comparing the treatment plans in the first two columns of Figure 3 (e.g., subjects 17904, 18582, and 19015), the 4 Gy isoline very closely follows the shape of any fMRI activations adjacent to the tumor mask whenever such a dose constraint is set on the fOARs, suggesting that the optimizer is successful in shaping the delivered dose in order to follow the dose constraints imposed on risk organs.

### 4.2. Effects of masking fMRI activation maps

We tested three masking settings with the goal of separating the closest fMRI activations from the tumor mask and thus improving the quality of the resulting treatment plans.

Removing all activations that fall inside the tumor results in a slight improvement in the treatment plan metrics, with a median target coverage of 0.83 and a median PCI of 0.73, respectively, while the median gradient index remains at 2.88. The median maximum dose decreases to 30.1 Gy, and the median minimum tumor dose increases to 2.65 Gy, indicating that the hot and cold spots in the treatment plan are somewhat alleviated. Although this constitutes a marked improvement for the treatment plans, most fMRI activations within the tumor mask do not appear deep inside the tumor, but are portions of larger activations adjacent to the tumor which extend into its edges (see Figure 3, second and third columns). The remaining portion of such activations, as well as any other activations immediately adjacent to the tumor, would shrink the portion of the tumor receiving the full treatment dose, and have a detrimental effect on the treatment plans.

Masking out all activations within 2 mm of all tumor voxels results in a further improvement to treatment plan metrics. Median target coverage and median PCI take values of 0.88 and 0.76, respectively, while the median gradient index remains nearly unchanged. Hot and cold spots are substantially reduced, with median maximum and minimum tumor doses of 3.85 Gy and 27.1 Gy, respectively. Increasing the masking radius to 4 mm provides an even larger improvement, with median target coverage and median PCI values of 0.98 and 0.82, respectively, and median maximum and minimum tumor doses of 22.65 Gy and 7.8 Gy, respectively. This brings most treatment plan metrics close to their values when not applying any dose limits, with the exception of the median gradient index, which remains at an elevated value of 2.88.

### 4.3. Optimization and treatment times

Both treatment plan optimization times and treatment times are affected by the inclusion of fOARs. Without fOARs the median time for generating a treatment plan is 0.45 minutes, or just under 30 seconds. The inclusion of fOARs increases the median optimization time for treatment plans to 5 minutes. Notably, optimization times are mostly dependent on the number and size of ROIs involved in the treatment planning, and are thus mostly unaffected by the various masking settings tested.

The median treatment time for patients when not using fOARs is 28 minutes, but with the inclusion of fOARs these times increase fourfold to 113 minutes. Much of this increase can be attributed to the presence of fMRI activation within and near the tumor mask, which give rise to complex treatment plans. While the masking of activations inside the tumors does not yield shorter treatment plans, the inclusion of 2 and 4 mm masking margins reduces median treatment times to 97 and 69 minutes, respectively.

### 4.4. Effects of spatial smoothing

Figure 4 presents a comparison of treatment plan metrics for plans where fOARs were produced using spatial filters of 4 mm and 6 mm FWHM. It should be noted that 6 mm filters gave rise to substantially more active voxels than 4 mm filters, with a median ratio of 3 (range 1.6 – 12.3). This is to be expected, as the use of larger filters increases the detection power of the analysis, but also introduces uncertainty over the exact location of activations. Nevertheless, most treatment plan metrics yield comparable results for both of the filter sizes tested, particularly when not setting dose limits on fOARs.

**Figure 4:**
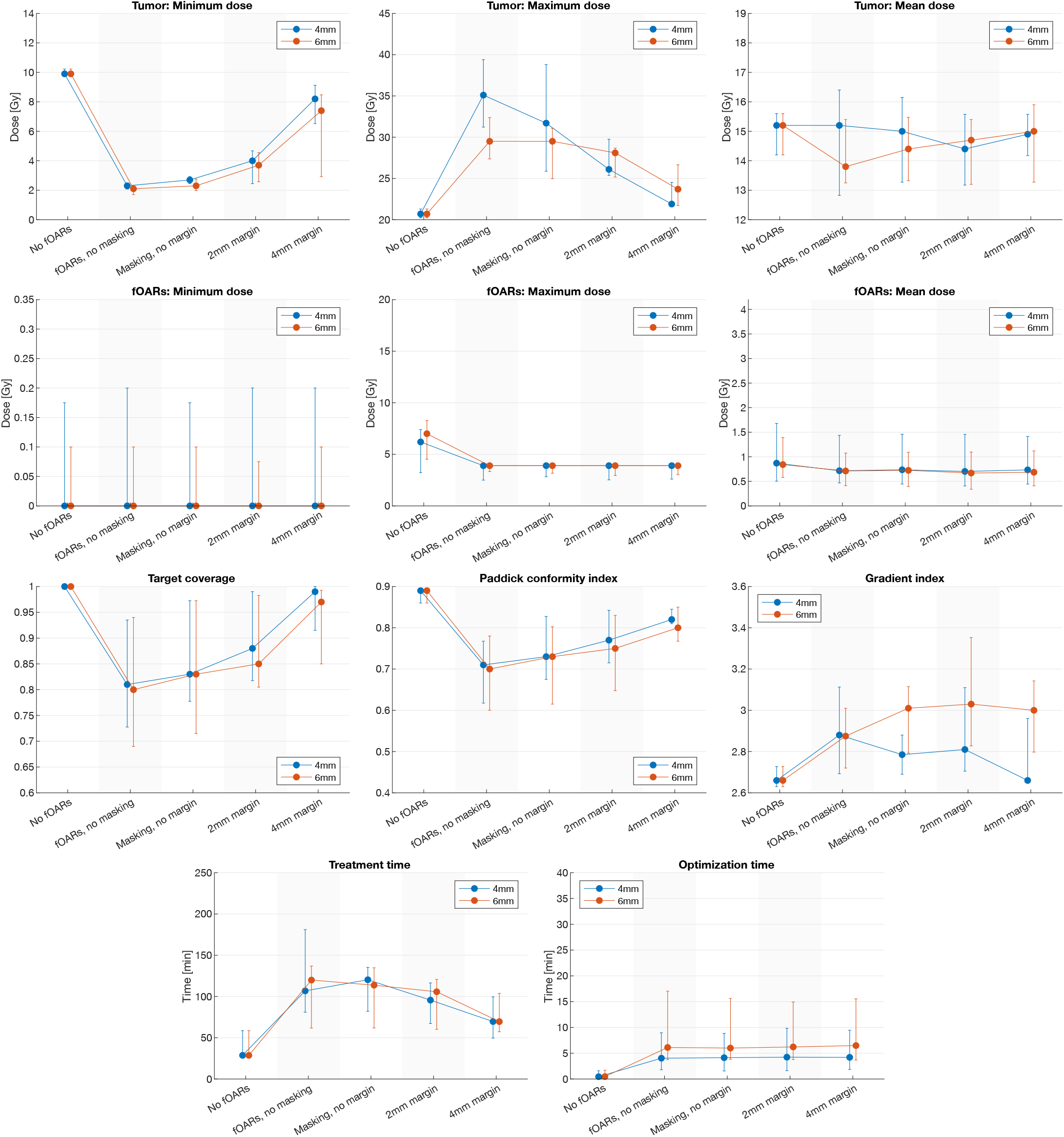
Comparison of effects of different spatial smoothing filter sizes on metrics for treatment plans generated using different fOAR dose constraints and masking settings. First plot row: tumor dose metrics. Second plot row: fOAR dose metrics. Third and fourth plot rows: treatment plan metrics. The first column inside each plot (“No fOARs”) shows results when no dose limits are set for the fOARs, while the remaining four columns represent the various masking settings employed. Dots and error bars represent the median and interquartile range respectively. Plot data ranges are over subjects.

When using fOARs, treatment quality metrics are consistently slightly worse for 6 mm filters than for 4 mm filters. Median target coverage and PCI are somewhat reduced for the 6 mm case, while median gradient indices are substantially higher than for 4 mm filters. These differences become more pronounced for larger margin sizes.

Comparison of tumor dose metrics is less straightforward. While the median minimum dose is slightly lower for 6 mm filters, the median maximum and mean doses are larger only for the cases when masking or margin are not used, while becoming lower than the ones for 4 mm filters when a masking radius is used.

For both filter sizes, median treatment plan optimization times when using fOARs are unaffected by masking and margin settings, and are shorter for 4 mm filters than for 6 mm filters, with medians of 4 and 6 minutes, respectively. Notably, median treatment times from the plans generated using both filter sizes are not substantially different.

### 4.5. Differences between subjects

Figure 5 presents individual treatment plan metrics obtained for each subject, illustrating that while there exists considerable variation in treatment plan quality at a subject level, the overall trends are shared by all subjects.

**Figure 5:**
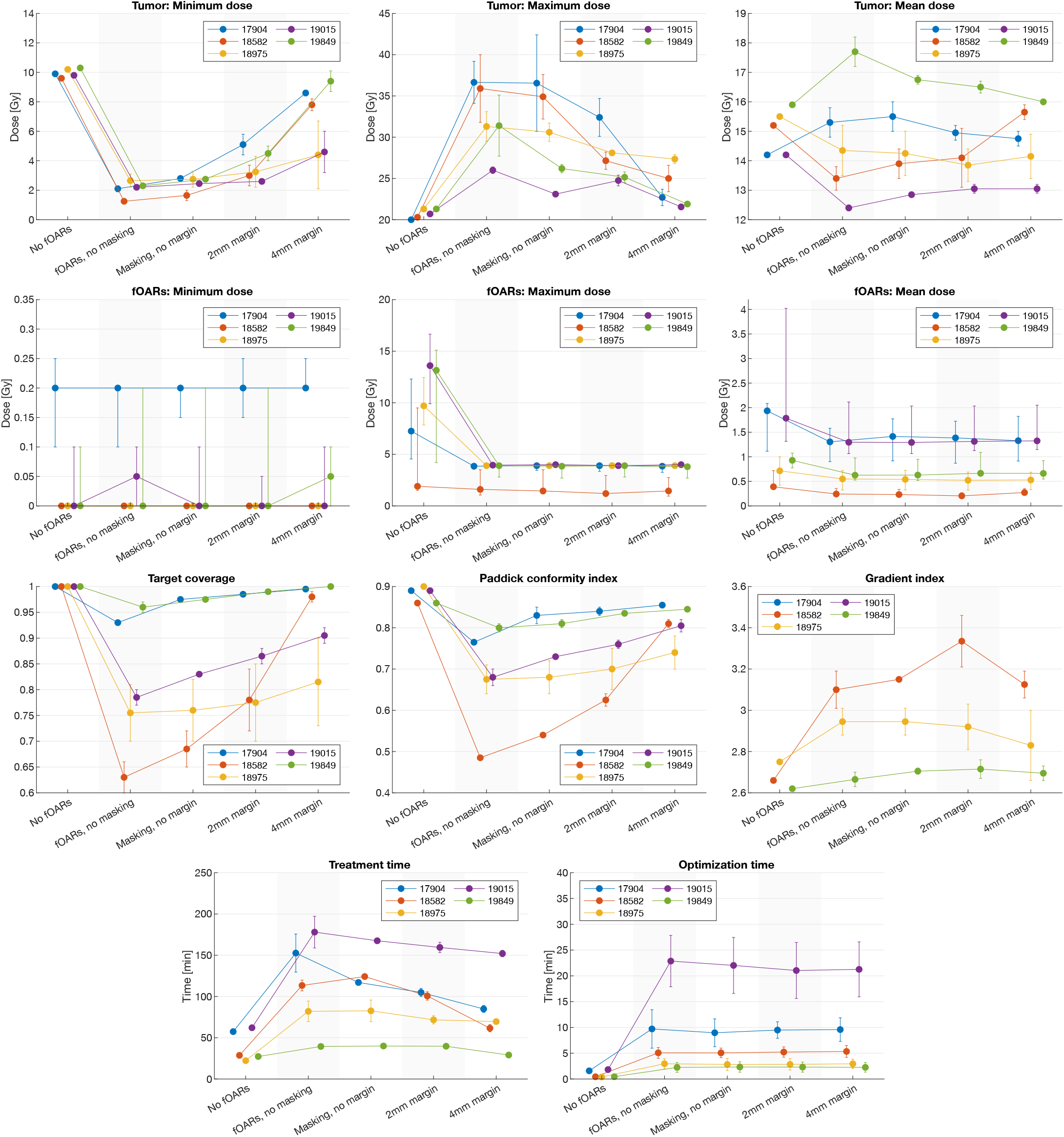
Comparison of individual subject metrics for treatment plans generated using different fOAR dose constraints and masking settings. First plot row: tumor dose metrics. Second plot row: fOAR dose metrics. Third and fourth plot rows: treatment plan metrics. The first column inside each plot (“No fOARs”) shows results when no dose limits are set for the fOARs, while the remaining four columns represent the various masking settings employed. Dots and error bars represent the median and interquartile range respectively. Plot data ranges are over smoothing filter sizes.

There is substantial subject-level variation in treatment quality metrics and in the relative benefits of applying masking and a margin on fOARs. For subjects 17904 and 19849, the inclusion of fOARs does not result in a substantial worsening of treatment plan quality, and masking bring quality metrics close to their values when no fOARs are applied. Conversely, for subjects 18975 and 19015 the use of fOARs results in a considerable reduction in treatment plan quality, and while masking provides a moderate improvement, quality metrics do not approach their values when fOARs are not used. Finally, subject 18582 is of particular interest, as they experience the greatest quality loss from the inclusion of fOARs, with target coverage and PCI being reduced by 0.37 and .38, respectively, yet disproportionately benefit from the use of masking, with the same metrics increasing by 0.35 and 0.33, respectively, when a 4 mm margin is used.

Tumor doses for different masking settings vary substantially across subjects. While subjects are differently affected by hot and cold spots, the use of masking and a margin around the tumor consistently reduces their presence for all subjects. Median maximum doses to fOARs for individual subjects can be as high as 13.6 Gy when dose constraints are not set, while the overall maximum dose observed on fOARs was 18.5 Gy. Nevertheless, setting dose constraints successfully limits the dose delivered to fOARs to 4 Gy or lower for all subjects.

Treatment plan optimization times show a strong dependence on tumor size, with larger tumors generally resulting in longer optimization times. Similarly, treatment times are are generally higher for subjects with larger tumors.

## 5. Discussion

Our results show that including fMRI-derived fOARs in the treatment planning process successfully limits the radiation dose incident on the eloquent brain regions, while still resulting in clinically acceptable treatment plans. In general, masking out the fOARs in a margin around the tumor results in treatment treatment plan metrics close to the ones obtained without including fOARs. In terms of radiation, our results are in line with previous work on using fOARs in treatment planning, showing that the eloquent brain areas receive substantially less radiation (Narayana et al., 2007; Kovács et al., 2011; Wang et al., 2015; Liu et al., 2000; Stancanello et al., 2007; Pantelis et al., 2010; Conti et al., 2013; De Martin et al., 2017).

Using the proposed method, it would be possible to incorporate fMRI-derived fOARs into the treatment planning protocol without requiring a large additional time investment. Collecting the fMRI data would require an additional 5–20 minutes in the MR scanner, depending on the number of tasks. While the preprocessing for a single subject can take approximately 100 minutes, most of this time is spent on preprocessing with fMRIPrep, and could be considerably reduced by using a different software tool or implementing a custom preprocessing pipeline.

From a user perspective, adding the fOARs in GammaPlan takes 1–3 minutes, while the optimization increases by 3–30 minutes, depending on the number and size of the targets and fOARs. From a treatment perspective, the treatment time increases from 20–60 minutes to 40-200 minutes, but this time can be substantially reduced to 20–160 minutes by the inclusion of a masking margin around the targets. In addition, the use of cluster extent thresholding of the fMRI activation maps, as well as stricter voxel-level thresholding, can potentially reduce the number of brain activity clusters, which would reduce the optimization and treatment times. It should also be noted that the Lightning planner can be tuned to reduce the treatment time, at the cost of increasing the dose outside the target.

### 5.1. Effects on patient quality of life

It is difficult to assess how using fOARs would affect the quality of life for the patients, as it would require a clinical study with and without fOARs in the treatment planning. Nevertheless, radiation induced damage to the healthy brain and its eloquent structures is a well-known and feared risk. Sequelae such as motor dysfunction, cognitive impairment, loss of hearing or vision, cranial nerve dysfunction or hypopituitarism all have a negative impact on patients. These unwanted side-effects are therefore sought to be avoided as far as reasonably possible while still maintaining sufficient treatment levels of radiation to the target tissue (Lambrecht et al., 2018). In this work we show that treatment dose of target tissue can be maintained with a reduced dose of radiation to vulnerable areas in the vicinity of the tumor, which should be beneficial to the patients treated.

### 5.2. Limitations

Preprocessing and statistical analysis of fMRI data presents a large number of degrees of freedom (Carp, 2012b,a). A non-exhaustive list of these includes the specific preprocessing steps performed (e.g., head motion correction, co-registration, denoising, slice timing correction, distortion correction), the specific methods use for each of these, the statistical methods used (e.g., voxel-level inference vs. cluster extent inference, Bayesian models, etc.). It would be very interesting to investigate how different preprocessing settings, and different statistical analyses, affect the generated treatment plans, but GammaPlan requires that each treatment plan is generated and exported manually. It is therefore not possible for us to make use any scripting tools to perform a large number of analyses by looping through different subjects and a large number of different settings. For this reason, we have limited the number of presented analyses in this paper. Without this limitation we could present results from many different settings for preprocessing and statistical analysis.

There is a degree of uncertainty inherent in the activation maps produced by fMRI. Activations frequently appear in regions other than expected from the task being performed, and may be disregarded. Furthermore, there is individual variability in the number, size, and location of activations (Bennett & Miller, 2010), which can exacerbated by patient pathologies (Silva et al., 2018). For these reasons, activation maps should be examined and thresholded by experienced neuroradiologists. This will however increase the planning time.

Stancanello et al. (2007) refer to two sources of error in their procedure for generating fOARs from fMRI. The first is the threshold set by expert neuroradiologists to determine the location and extent of cortical activations. The second is the cumulative spatial imprecision resulting from chaining multiple registration steps. To this we must add that the data we used lacked any means for correcting susceptibility-based distortion. Although distortion correction without fieldmaps (Esteban et al., 2019) has been shown to produce better result than no distortion correction, it is an experimental methodology, and fieldmap-based correction would have been preferred. In our case the registration between the fMRI data and the T1w volume will therefore not be perfect.

Our work was carried out on a dataset where structural MRI was acquired without gadolinium contrast, making it more difficult to clearly delineate tumor boundaries. However, as this work is a proof-of-concept where no actual patients were treated, the precise delineation of tumor borders is not essential.

### 5.3. Experiences with GammaPlan

In our initial experiments with GammaPlan, each full activation map was treated as a single ROI, despite being composed of multiple unconnected components (clusters), as the RTSTRUCT format imposes no constraints on ROIs having to be single connected components. However, since GammaPlan implicitly creates a single connected volume as a triangulated mesh based on all contours belonging to the same ROI, such combined ROIs gave way to surface artifacts in the form of spurious connections and self-intersections. Instead, it is necessary to treat each individual connected component of the activation maps as a separate ROI, which was done in all reported experiments. However, this dramatically increases the number of ROIs that need to be managed in GammaPlan (up to anywhere from several dozen to almost two hundred in our case). This can be inconvenient, as for each ROI it is necessary to specify its type, i.e. target or region of avoidance, with its corresponding target or maximum dose, respectively. While the ROI type can be specified in the DICOM metadata before loading them into GammaPlan, the dose settings have to be set manually by the GammaPlan operator for each ROI.

Although treating each activation cluster as a single ROI removed most artifacts in GammaPlan, there were some remaining problems in approximately half of the 50 generated treatment plans. The imported 2D contours for fOARs look correct in the axial plane, but self intersections are visible in the coronal and sagittal planes. The 3D surface generated from these contours, used by the optimizer for dose calculations, occasionally intersects the tumor mask, resulting in incomplete target coverage and suboptimal treatment plans. This problem was present even in cases where fOARs where masked out within a radius around the tumor targets, which should preclude any overlap between fOARs and the tumor mask. As an independent validation of our workflow, we imported the same DICOM and RTSTRUCT files into the 3D Slicer software^3^, and observed that these problems were not present. The fact that the surface reconstruction in GammaPlan causes intersections between some fOARs and the tumor masks results in impaired treatment plans, and we believe that some of the reported treatment metrics would be further improved without this problem.

### 5.4. Clinical usage of fMRI in SRS

At Linköping University hospital, fMRI is part of the clinical protocol for brain tumor resection surgery in relevant cases, but not for SRS. There are at least two reasons why fMRI is not commonly used to generate fOARs for SRS. Firstly, automatic inverse planners have only recently become available for Gamma Knife SRS planning. To manually create 50 treatment plans while considering 80–180 fOARs for each would be prohibitively time-consuming. Secondly, creating a pipeline that analyzes the fMRI data and generates RTSTRUCT files was a non-trivial task. For instance, the conversion of data from NIfTI to DICOM format is substantially more involved than the opposite conversion. RTSTRUCT and DICOM are rather complex file formats, whereas neuroimaging researchers are mostly used to the simpler NIfTI file format. Our hope is that this work will increase the use of fOARs in SRS, and that our shared code can help other researchers and clinicians who want to continue our work.

One factor that could simplify the incorporation of fMRI data into clinical tumor treatment protocols is the adoption of DICOM Segmentation objects as a potential data format for the definition of dose optimization constraints. DICOM Segmentation objects provide a means for associating binary or fractional classification volumes to reference DICOM images, which makes them ideally suited for storing brain activation maps in their native form, obviating the need for the cumbersome conversion step of activation maps into collections of contours required when using DICOM RT Structure Sets. Furthermore, the capacity to represent both binary and fractional classification volumes would allow them to store both thresholded and unthresholded activity maps, respectively. While DICOM RT Structure Sets are well suited for representing manual organ delineations, DICOM Segmentation objects are a better fit for representing the output of MRI processing pipelines and machine-generated organ delineations.

## 6. Conclusion

Reducing the amount of gamma radiation incident on the eloquent regions of the brain can help minimize the risk of neurological complications after SRS, and improve the quality of life of patients. Task-based fMRI provides a way for localizing these regions. We have demonstrated that fMRI-derived fOARs can be incorporated into Gamma Knife SRS treatment planning to minimize the radiation dose incident on the eloquent regions of the brain, while maintaining acceptable treatment quality metrics. Although we have focused on task-based fMRI, similar pipelines can be set up to generate OARs from other MRI modalities, such as resting state fMRI and diffusion MRI, with the goal of reducing the radiation incident on specific brain networks and fiber tracts, respectively.

## Data Availability

This work used open data published in Pernet et al., 2016, "A structural and functional magnetic resonance imaging dataset of brain tumour patients".

## Ethics

The ethical review board of Linköping decided that no additional ethical approval is required to analyze the openly available fMRI dataset.

## Acknowledgements

This study was performed in collaboration with Elekta Instrument AB (Stockholm, Sweden), and the authors are grateful for the opportunity of using the latest research version of Elekta Leksell GammaPlan which includes the inverse planner Lightning. We thank Håkan Nordström, Jonas Johansson, Björn Somell, Stella Riad and Kenneth Lau for sharing their valuable knowledge about GammaPlan. We thank Cyril Pernet et al. for collecting and sharing the used dataset.

This study was supported by CENIIT at Linköping University, by the ITEA3 / VINNOVA funded project “Intelligence based iMprovement of Personalized treatment And Clinical workflow supporT” (IMPACT), by LiU cancer at Linköping University, and by the ITEA3 / VINNOVA funded project “Automation, Surgery Support and Intuitive 3D visualization to optimize workflow in IGT SysTems” (ASSIST). The authors did not receive any funding from Elekta Instrument AB.

## Appendix A. fMRIPrep preprocessing

The exact function call used for fMRIPrep was `fmriprep --random-seed 100 --skull-strip-t1w auto --output-spaces T1w --use-syn-sdc --fs-no-reconall --fs-license-file /license.txt /data /out participant`.

Following is the detailed description of the preprocessing steps carried out by fMRIPrep. This description was itself automatically generated by fMRIPrep:

Results included in this manuscript come from preprocessing performed using *fMRIPrep* 21.0.0 (Esteban et al., 2019, 2018, RRID:SCR 016216), which is based on *Nipype* 1.6.1 (Gorgolewski et al., 2011, 2018, RRID:SCR 002502).

### Appendix A.1. Anatomical data preprocessing

A total of 1 T1-weighted (T1w) images were found within the input BIDS dataset. The T1-weighted (T1w) image was corrected for intensity non-uniformity (INU) with `N4BiasFieldCorrection` (Tustison et al., 2010), distributed with ANTs 2.3.3 (Avants et al., 2008, RRID:SCR 004757), and used as T1w-reference throughout the workflow. The T1w-reference was then skull-stripped with a *Nipype* implementation of the `antsBrainExtraction.sh` workflow (from ANTs), using OASIS30ANTs as target template. Brain tissue segmentation of cerebrospinal fluid (CSF), white-matter (WM) and gray-matter (GM) was performed on the brain-extracted T1w using `fast` (FSL 6.0.5.1:57b01774, RRID:SCR 002823, Zhang et al., 2001). Volume-based spatial normalization to one standard space (MNI152NLin2009cAsym) was performed through nonlinear registration with `antsRegistration` (ANTs 2.3.3), using brain-extracted versions of both T1w reference and the T1w template. The following template was selected for spatial normalization: *ICBM 152 Nonlinear Asymmetrical template version 2009c* (Fonov et al., 2009, RRID:SCR 008796; TemplateFlow ID: MNI152NLin2009cAsym).

### Appendix A.2. Preprocessing of B0 inhomogeneity mappings

A total of 1 fieldmaps were found available within the input BIDS structure for this particular subject. A deformation field to correct for susceptibility distortions was estimated based on *fMRIPrep*’s *fieldmap-less* approach. The deformation field is that resulting from co-registering the EPI reference to the same-subject T1w-reference with its intensity inverted (Wang et al., 2017; Huntenburg, 2014). Registration is performed with `antsRegistration` (ANTs 2.3.3), and the process regularized by constraining deformation to be nonzero only along the phase-encoding direction, and modulated with an average fieldmap template (Treiber et al., 2016).

### Appendix A.3. Functional data preprocessing

For each of the 2 BOLD runs found per subject (across all tasks and sessions), the following preprocessing was performed. First, a reference volume and its skull-stripped version were generated using a custom methodology of *fMRIPrep*. Head-motion parameters with respect to the BOLD reference (transformation matrices, and six corresponding rotation and translation parameters) are estimated before any spatiotemporal filtering using `mcflirt` (FSL 6.0.5.1:57b01774, Jenkinson et al., 2002). The estimated *fieldmap* was then aligned with rigid-registration to the target EPI (echo-planar imaging) reference run. The field coefficients were mapped on to the reference EPI using the transform. The BOLD reference was then co-registered to the T1w reference using `mri coreg` (FreeSurfer) followed by `flirt` (FSL 6.0.5.1:57b01774, Jenkinson & Smith, 2001) with the boundary-based registration (Greve & Fischl, 2009) cost-function. Co-registration was configured with six degrees of freedom. Several confounding time-series were calculated based on the *preprocessed BOLD* : framewise displacement (FD), DVARS and three region-wise global signals. FD was computed using two formulations following Power (absolute sum of relative motions, Power et al. (2014)) and Jenkinson (relative root mean square displacement between affines, Jenkinson et al. (2002)).

FD and DVARS are calculated for each functional run, both using their implementations in *Nipype* (following the definitions by Power et al., 2014). The three global signals are extracted within the CSF, the WM, and the whole-brain masks. Additionally, a set of physiological regressors were extracted to allow for component-based noise correction (*CompCor*, Behzadi et al., 2007). Principal components are estimated after high-pass filtering the *preprocessed BOLD* time-series (using a discrete cosine filter with 128s cut-off) for the two *CompCor* variants: temporal (tCompCor) and anatomical (aCompCor). tCompCor components are then calculated from the top 2% variable voxels within the brain mask. For aCompCor, three probabilistic masks (CSF, WM and combined CSF+WM) are generated in anatomical space. The implementation differs from that of Behzadi et al. in that instead of eroding the masks by 2 pixels on BOLD space, the aCompCor masks are subtracted a mask of pixels that likely contain a volume fraction of GM. This mask is obtained by thresholding the corresponding partial volume map at 0.05, and it ensures components are not extracted from voxels containing a minimal fraction of GM. Finally, these masks are resampled into BOLD space and binarized by thresholding at 0.99 (as in the original implementation). Components are also calculated separately within the WM and CSF masks.

For each CompCor decomposition, the *k* components with the largest singular values are retained, such that the retained components’ time series are sufficient to explain 50 percent of variance across the nuisance mask (CSF, WM, combined, or temporal). The remaining components are dropped from consideration. The head-motion estimates calculated in the correction step were also placed within the corresponding confounds file. The confound time series derived from head motion estimates and global signals were expanded with the inclusion of temporal derivatives and quadratic terms for each (Satterthwaite et al., 2013). Frames that exceeded a threshold of 0.5 mm FD or 1.5 standardised DVARS were annotated as motion outliers. All resamplings can be performed with *a single interpolation step* by composing all the pertinent transformations (i.e. head-motion transform matrices, susceptibility distortion correction when available, and co-registrations to anatomical and output spaces). Gridded (volumetric) resamplings were performed using `antsApplyTransforms` (ANTs), configured with Lanczos interpolation to minimize the smoothing effects of other kernels (Lanczos, 1964). Non-gridded (surface) resamplings were performed using `mri vol2surf` (FreeSurfer).

Many internal operations of *fMRIPrep* use *Nilearn* 0.8.1 (Abraham et al., 2014, RRID:SCR 001362), mostly within the functional processing workflow. For more details of the pipeline, see [the section corresponding to workflows in *fMRIPrep*’s documentation](https://fmriprep.readthedocs.io/en/latest/workflows.html).

## Footnotes

1 https://github.com/DavidAbramian/CENIIT2

2 software build: alpha1A1-nonclinical (6576.d71bbebac) @ sesrdtpsbuild005 2019-10-10T11:55

3 https://www.slicer.org

